# Spatial Modeling of Sociodemographic Risk for COVID-19 Mortality

**DOI:** 10.1101/2023.07.21.23292785

**Authors:** Erich Seamon, Benjamin J. Ridenhour, Craig R. Miller, Jennifer Johnson-Leung

## Abstract

In early 2020, the Coronavirus Disease 19 (COVID-19) rapidly spread across the United States (US), exhibiting significant geographic variability. While several studies have examined the predictive relationships of differing factors on COVID-19 deaths, few have looked at spatiotemporal variation at refined geographic scales. The objective of this analysis is to examine this spatiotemporal variation in COVID-19 deaths with respect to association with socioeconomic, health, demographic, and political factors. We use multivariate regression applied to Health and Human Services (HHS) regions as well as nationwide county-level geographically weighted random forest (GWRF) models. Analyses were performed on data from three separate time frames which correspond to the spread of distinct viral variants in the US: pandemic onset until May 2021, May 2021 through November 2021, and December 2021 until April 2022. Multivariate regression results for all regions across three time windows suggest that existing measures of social vulnerability for disaster preparedness (SVI) are predictive of a higher degree of mortality from COVID-19. In comparison, GWRF models provide a more robust evaluation of feature importance and prediction, exposing the value of local features for prediction, such as obesity, which is obscured by coarse-grained analysis. Overall, GWRF results indicate that this more nuanced modeling strategy is useful for determining the spatial variation in the importance of sociodemographic risk factors for predicting COVID-19 mortality.

## 1. Introduction

The burden of COVID-19 in the United States from the onset of the pandemic through early 2022 was unevenly distributed throughout the population[1,2]. The elderly were identified as being particularly vulnerable to severe disease and death from the outset of the pandemic [3,4]. Co-morbid conditions such as obesity, diabetes, heart disease, and hypertension are also significant risk factors for COVID-19 mortality [5,6]. However, these observations are insufficient to explain the wide variation in COVID-19 burden. In this study, we analyze the importance of social vulnerability, demographic and health parameters, and political geography in predicting COVID-19 county-level death rates in the United States. Specifically, we use a spatially-explicit modeling technique in order to improve the accuracy of prediction at a geographically fine scale.

A potential explanation for the extreme variability in COVID-19 burden across the U.S. is the high variability of social risk factors within the population [7–9], with one of the most notable social divides being the urban/rural disparity in the U.S. In particular, rural communities have been identified by the World Health Organization (WHO) as being disproportionately elderly and having fewer healthcare resources [10]. Similarly, these communities have increased rates of poverty, with 15.4 percent of rural populations living below the federal poverty line, versus 11.9 percent in urban areas [11]. The U.S. urban/rural disparity is further evidenced by examining insurance access [12], telecommunications/broadband access [13], housing [14], and political ideology [15]. We hypothesize that modeling these social factors using cutting-edge fine-scale spatial models provides a richer, more nuanced understanding of the association of these disparities with COVID-19 mortality.

Further complicating the epidemiology of COVID-19, political ideology has been identified as an unexpectedly effective predictor for COVID-19 mortality [16]. Given the importance of trust [17] in public health messengers in driving protective behavior, politicization of the issue is particularly problematic. Specifically, political ideology has been tied to the adoption of protective behaviors (i.e., masking, social distancing, vaccination) and created a patchwork of federal, state, and local policies [18] combined with individual responses to these policies [17]. The importance of political ideology may be explained in part by the ability of groups to recruit followers and refine messaging via online platforms; this ability to create “echo chambers” has increased due to the wide adoption of social media and the increase of remote work, school, and social activities[19]. If group messaging reinforces actions counter to recommended health behavior, populations which are most at risk bear the brunt of these consequences (i.e., rural communities). We therefore include measures of political leaning in our models.

Other researchers have also attempted to use spatial models to predict COVID-19 outcomes. For example, Anderson et al. [20] used cluster analysis to evaluate spatial patterns of transmissibility and their relationships to social health parameters. They later combined this approach with a three-stage regression technique, to explore impacts to mortality and morbidity [21]. In another work, Sun et al. [22] looked at spatial lag aspects of COVID-19 in the United States, compared to county-level demographic variables. Similarly, Mollalo et al. [23,24] evaluated differing regression-based approaches to examine spatial heterogeneity, while Xie et al. [25] applied exploratory spatial analysis methods to find associations with demographic and social variables. A common theme for all of the above research efforts were the difficulties in optimizing spatial interactions, in combination with time scales. To address these concerns, we use a modeling strategy (Figure 1) which compares more traditional (linear) regression to geographically weighted random forests (GWRF), and study the spatial autocorrelation of these factors using a Moran’s I analysis.

**Figure 1.**
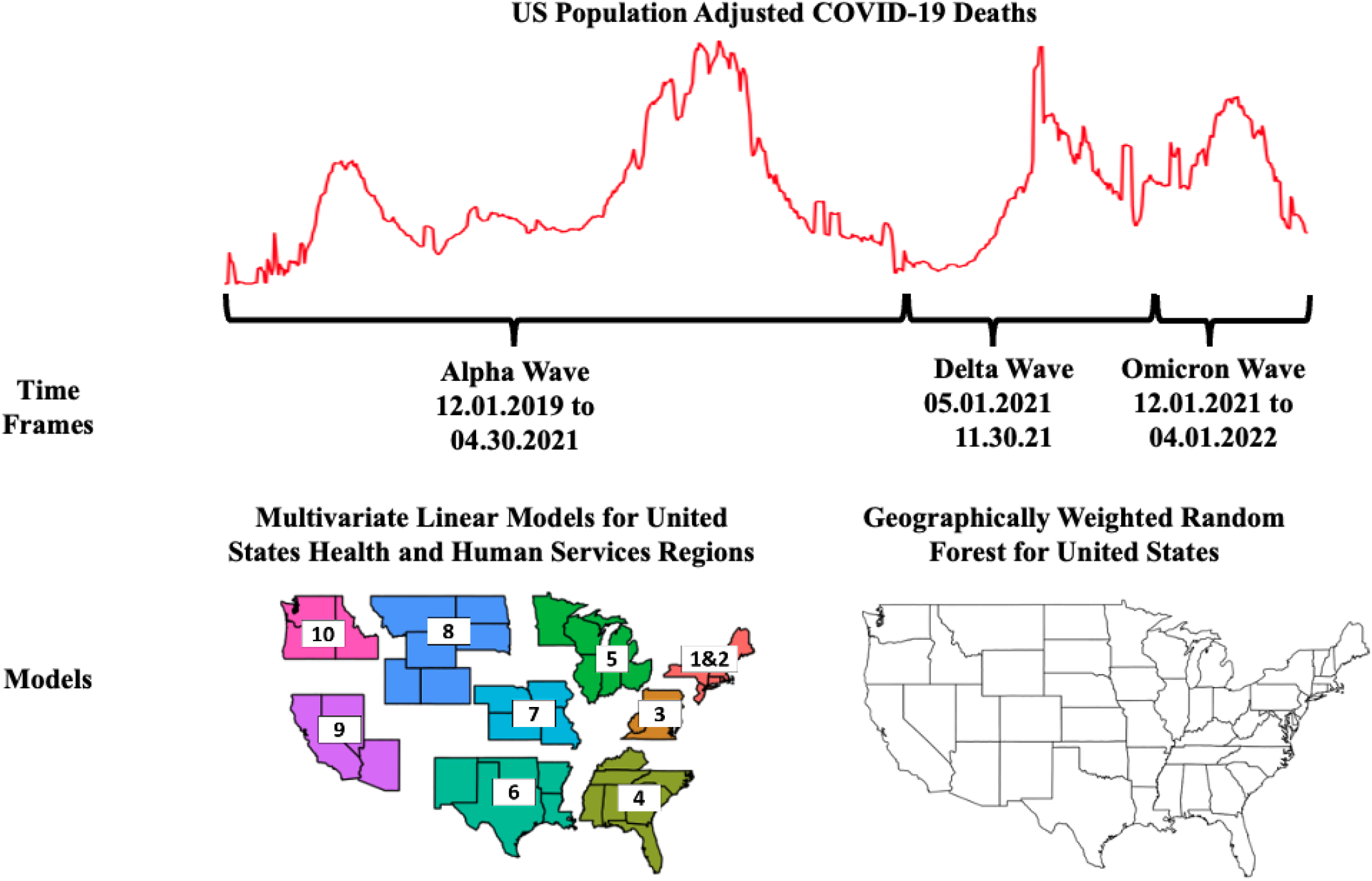
Analysis scheme. Our approach examines fifteen (15) different county-level independent variables, in relationship to cumulative population adjusted deaths during a pandemic wave. We regress our variables at the regional level, and subsequently use them for spatially weighted random forest models. In total, thirty-three (33) models were run (a national regression model, nine regional regression models, and a national GWRF model for each of the three time periods). We also evaluate spatial heterogeneity at a county level by using local and global Moran’s I values, with Monte Carlo simulations performed to minimize error [30,31].

A GWRF is a statistical model that combines the strengths of two well-established techniques: Geographically Weighted Regression (GWR) and Random Forests (RF). GWR is a type of spatial regression model that allows the estimation of regression coefficients that vary spatially. This permits the model to capture local patterns in the data, which is particularly useful when dealing with data that exhibit spatial heterogeneity [26]. However, GWR is limited in its ability to handle complex nonlinear relationships between variables. Comparatively, a RF is an ensemble machine learning model that can handle complex non-linear relationships between variables and has the ability to capture variable interactions [27]; RF models are a popular choice for large data sets due to low computational costs. The GWRF modeling approach combines the strengths of GWR and RF by weighting the RF model locally, allowing it to capture local patterns in the data while also handling complex nonlinear relationships [28]. While GWRF modeling is not suited for hypothesis testing or causal inference, the method has the ability to handle spatial heterogeneity in data and to provide predictions that are locally accurate. The value of the GWRF approach lies in its ability to provide accurate and locally relevant predictions for complex spatial data, and is particularly useful for environmental and geographical applications where spatial heterogeneity is a major concern, such as the COVID-19 pandemic data [29].

## 2. Materials and Methods

We initially performed exploratory data analyses of fatality rates and deaths across time, for the entire United States, as well as by region. Based on this review of the data, we developed a set of modeling frameworks for three time frames, approximately corresponding to the Alpha, Delta, and Omicron waves in the United States. For our analyses, we define: the Alpha wave as the period from the beginning of the pandemic through the universal availability of COVID-19 vaccines to adults in the United States (January 2020–April 2021); the Delta wave as the period from May 2021–November 2021; and lastly, the Omicron wave as the shortest period December 2021–April 2022. The response variable in all of our analyses is the cumulative US county-level COVID-19 deaths, adjusted for population, for the full time period of interest. Fifteen (15) predictor variables are evaluated as county-level measures: socioeconomic status, household composition and disability, minority status and language, housing type and transportation, voting percentage, vaccination rate estimates as of April 2022, population density, obesity, unemployment, uninsured adults, social associations, diabetes, food insecurity, broadband access, and the percentage of population over 65. Correlations among these variables are provided in Table S4 of the supplemental appendix.

We first evaluate spatial autocorrelation of all variables, for all three time frames, using Moran’s I. We then use two different modeling approaches in our analyses. The first approach uses multilinear Poisson regression for nine (9) regional areas of the United States, plus the entire United States as a singular model, for a total of ten (10) distinct county groupings. Regional models are based on Health and Human Services (HHS) divisions, which were selected as a generalized policy/funding separation (HHS, http://hhs.gov). Regions 1 and 2 are combined into one region because of the small number of states in Region 2. The second approach is a more novel geographically weighted random forest modeling technique for the United States as a whole. Each model is evaluated for each of the pandemic time periods, for a total of thirty-three (33) models (Fig.1). The full set of these analyses can be seen in the associated supplemental materials.

### Data Assembly

Predictor data are collected from a number of sources: variable descriptions and sources are listed in Table 1. All data were normalized based on a 0 to 1 scaling structure. Figure 2 shows the spatial distributions of a select set of independent variables. Of particular note are issues of missing data related to vaccinations. Early in the pandemic, a number of states (Texas, New Hampshire) failed to report vaccination data: as such, assessing vaccination rates early on (2020) is not possible for the entire United States. Because of this, we use county-level vaccination rates (defined as the percentage of individuals receiving at least one dose of a vaccine) as of April 2022; thus our vaccination measure reflects the portion of the population willing to (eventually) seek out a vaccine, rather than the portion of the population actually immunized at a particular time. Additionally, a number of sparsely populated counties (<20) have reporting errors with regards to deaths and case counts. In order to appropriately fit our models, these counties are excluded from our analyses.

**Table 1.**
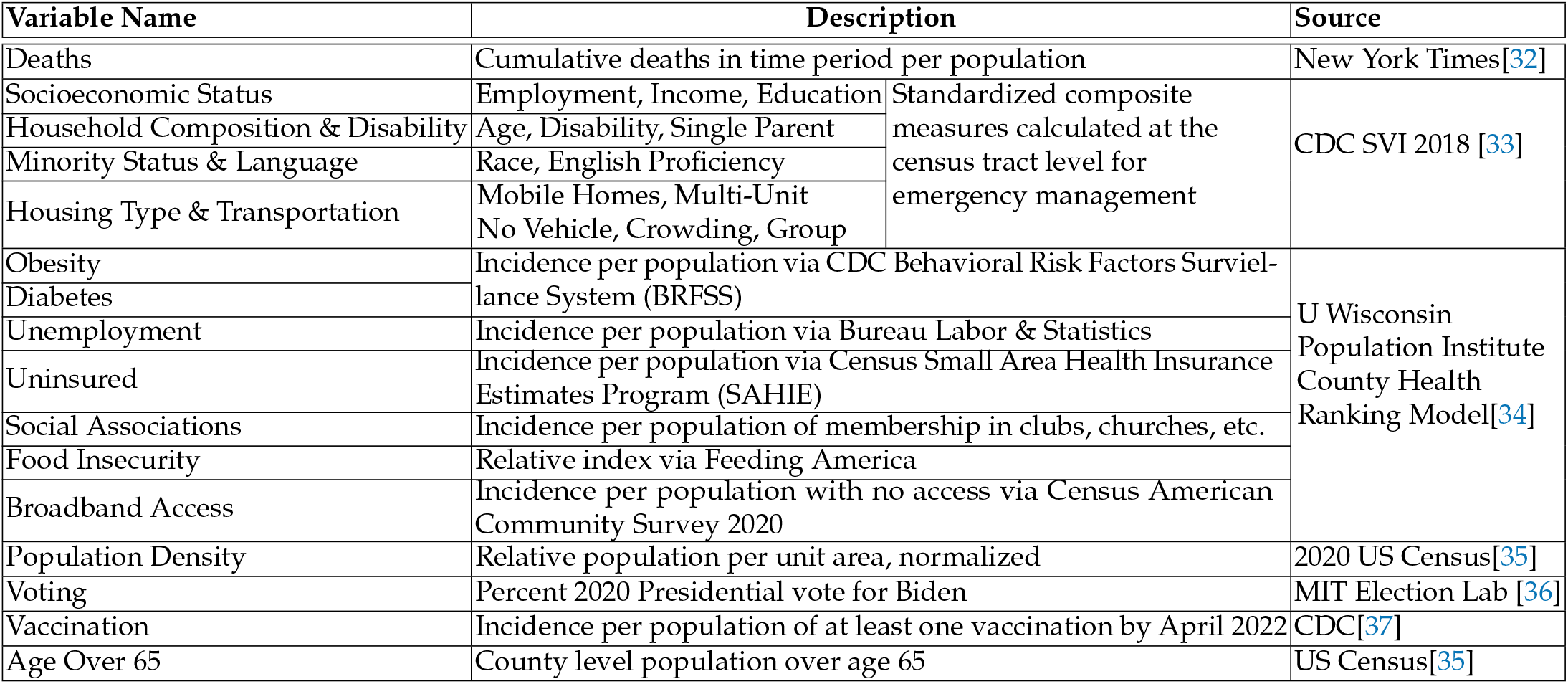
Description and Sources of Model Variables.

**Figure 2.**
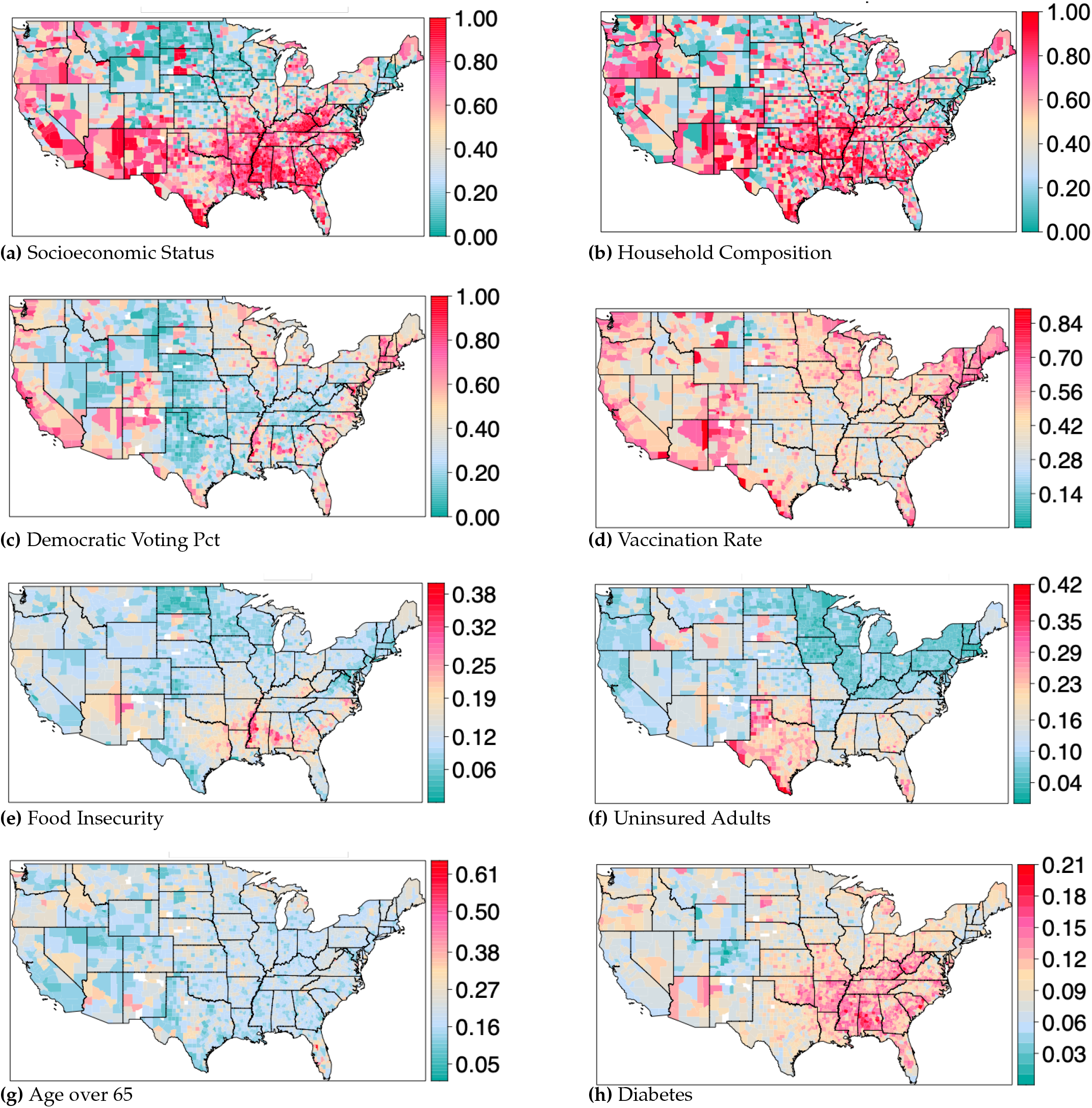
Select predictor variables mapped for the entire United States. a) Socioeconomic status: SVI representing income, poverty, employment, and education. b) Household composition: SVI representing levels of single parent households, disabilities, or those with children or the elderly. c) Democratic voting percentage: Percentage of democratic vote from the 2020 general presidential election. d) Vaccination rate: Percentage of individuals in a county with at least one vaccine dose, as of April 1, 2022. e) Food insecurity: Percentage of households with insufficient access to food, or food of an adequate quality. f) Uninsured adults: Percentage of adults that are uninsured. g) Age over 65: Percentage of individuals over age 65. h) Diabetes. Percentage of adults that have diabetes. Maps for all fifteen predictor variables can be found in the associated supplemental materials.

### Spatial Autocorrelation

The Moran’s I statistic,

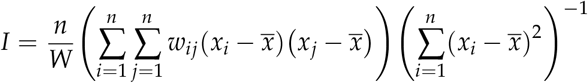

measures spatial autocorrelation[30,31,38]. Here *n* is the number of spatial units, *w*_*ij*_ are spatial weights, *x* is the variable being tested for autocorrelation with mean 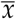, and *W* =∑_*i,j*_ *w*_*ij*_. The Moran’s I weight matrix specifies the degree of spatial proximity between pairs of spatial units, and can be calculated using contiguity-based weighting, network-based weighting, or distance-based weighting. Here we use contiguity-based weighting, which can be described as a n × n positive symmetric matrix W, where the element of this matrix is *w*_*ij*_ at location *i, j* for *n* locations. There are a number of contiguity weight approaches, including linear contiguity, rook contiguity, bishop contiguity, and queen contiguity [39]. Here we use queen contiguity, which creates a neighborhood list based on a common edge or a common vertex. Queen contiguity is recommended when irregular polygons (e.g. counties) are used [40]. This provides a method to evaluate spatial patterns in terms of positive (clustering), negative (dispersion), or neutral (random) spatial autocorrelation, with a range from -1 to 1. By examining Local Moran’s I values for each observation (in this instance, an individual county), we can look at clustering in combination with other factors (e.g. voting and mortality). Such comparisons across differing time frames give insight into how COVID-19 deaths may be spatially correlating with covariates [41].

### Regression Analysis and Geographically Weighted Random Forest

We construct Poisson regression models applied to HHS regions (with population as an offset) for each of the three time windows (Alpha, Delta, and Omicron), using all fifteen variables, with cumulative county-level death counts as the dependent variable. Normality assumptions are evaluated, including residual and partial residual/component residual plots, as well as standardized beta coefficients.

To address some of the limitations of the regression analysis, a modified novel approach of ensembled regression decision trees (random forest) were used. Regression decision trees are a method of constructing a set of decision rules on a predictor variable [42–44] that is continuous (versus categorical). These rules are constructed by recursively partitioning the data into successively smaller groups with binary splits based on a single predictor variable, with the goal of encapsulating the training data in the smallest possible tree [45]. Random forest, or ensembled decision trees, are a combination of many decision tree predictors, where each tree depends on the values of a random vector, sampled independently and with the same distribution for all trees in the forest [27]. Random forest modeling reduces the potential for over-fitting through the use of bootstrap aggregation, averaging across many trees, and provides a level of feature importance for assessing predictor power[42,46].

Geographically weighted random forest (GWRF) is a modified version of the classic random forest approach, which incorporates spatial non-stationarity, as part of spatially weighted regression (SWR) techniques [28]. GWRF fits a sub-model for each observation in space, taking into account neighboring observations (in this instance, observations are represented as counties), and is based on Fotheringham, Yang and Kang’s [26] work on spatially-explicit coefficient modeling. The main difference between a traditional (linear) SWR and GWRF is that we can model non-stationarity within a non-linear model, which minimizes over-fitting and thus relaxes the assumptions of traditional Gaussian statistics. As part of our GWRF construction, we utilized 10-fold cross validation, a model validation technique used to assess the generalizability of the model. Model construction for this analysis used the recursive partitioning and regression trees package (*rpart*), as well as the *gfr* package within R [42,46]. For the three sets of models developed (Alpha, Delta, and Omicron wave time periods), external cross validation was performed, using an adaptive kernel distance weighting function across a range of bandwidth values (number of surrounding counties, which ranged from 3 to 30).

The adaptive kernel function as part of the GWRF model is defined as:

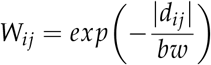

where *w*_*ij*_ is the weight assigned to the observation *j* for the estimation of *i*, and *d*_*ij*_ is the distance between *j* and *i*, and *bw* is the bandwidth (number of surrounding counties). Each variant model was run for progressively larger bandwidths, in combination with inversely varying global and local model weighting (local weighting at .25, .50, .75, and 1, combined with global weighting at 1, .75, .50, .25), to evaluate both root mean square error (RMSE) as well as mean absolute errors (MAE). Using this optimization method, we were able to select the model parameters which minimized error for each variant window (Supplemental Figures S23, S27, S31).

## 3. Results

All timeframes (Alpha, Delta and Omicron) indicated an overall positive spatial clustering, with moderate outliers, for COVID-19 deaths (localized Moran’s I, M = 0.462). By plotting deaths vs spatially lagged values (Fig. 3a) we see a progression from negative spatial autocorrelation for more liberal counties (blue counties, lower left quadrant) to positive spatial autocorrelation for more conservative counties (red counties, upper right quadrant). Outliers were overwhelmingly conservative-voting counties, which suggests that conservative counties are having a stronger influence over model fit. In addition, Monte Carlo simulations confirmed strong positive spatial autocorrelation (spatial clustering), with a density peak to the left of mean cumulative deaths values, for all three time windows. When examining the clustering behavior for independent variables, we see varying degrees of positive spatial clustering, as measured by the Moran’s I statistic. Fig. 3b shows that household composition is spatially correlated and that increased death rates were observed in poorer counties. Similar effects can be seen for the other SVI measures (Fig. 3c) as well as known health risk factors, such as diabetes (Fig. 3d) A number of variables—uninsured adults, social associations, unemployment—show no consistent pattern of deaths in relation to spatial clustering. See Part 4 of the supplementary materials for Moran’s scatterplots for each of the variables in all three waves.

**Figure 3.**
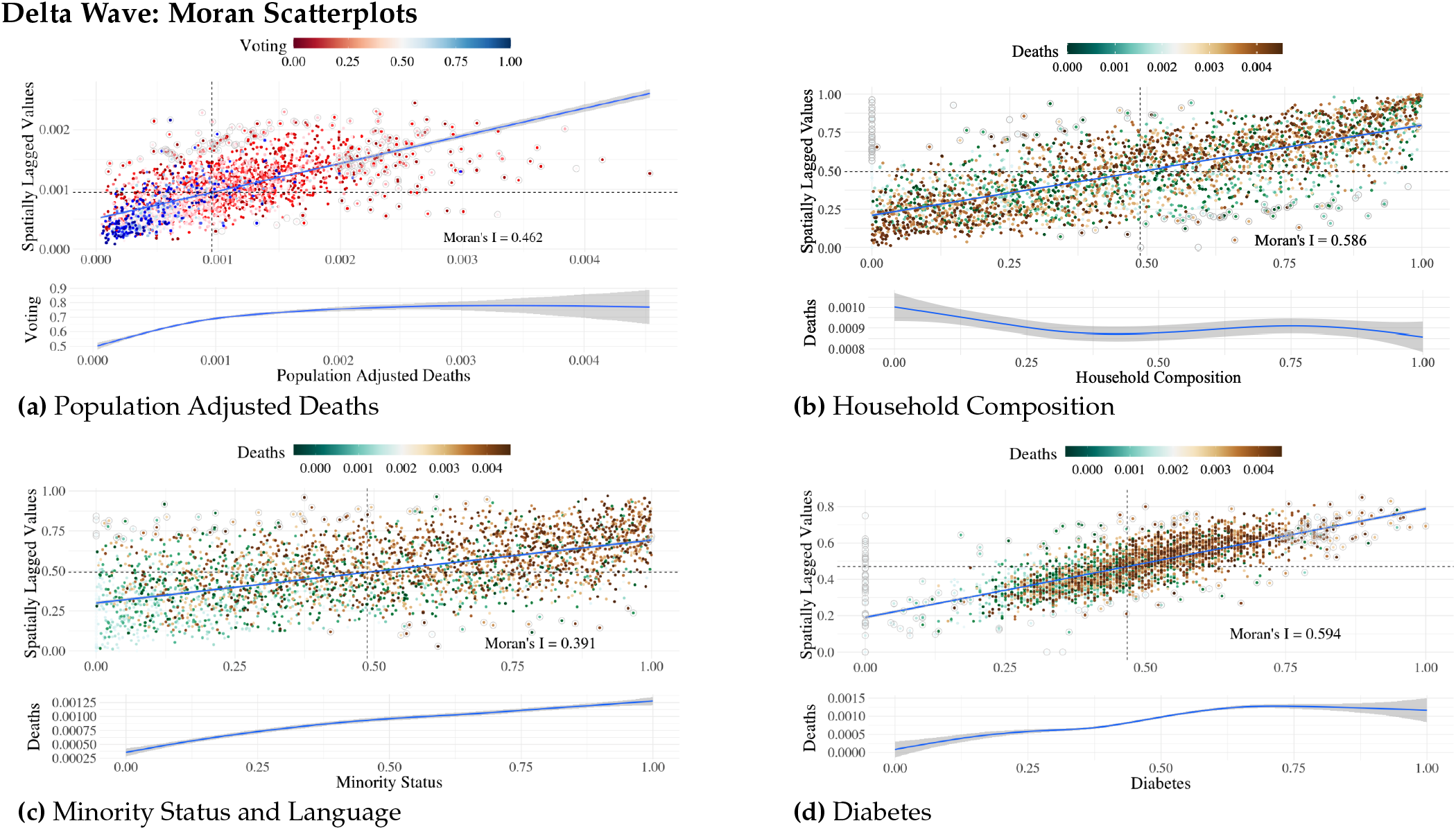
Select spatial autocorrelation Moran’s scatterplots for the Delta wave time window. Each point represents an individual county value. a) COVID-19 Population adjusted deaths Moran’s scatterplot. Color range depicts voting. b) Household Composition Moran’s scatterplot. Color range depicts COVID-19 population adjusted deaths. c) Minority Status and Language Moran’s scatterplot. Color range depicts COVID-19 population adjusted deaths. d) Diabetes Moran’s scatterplot. Color range depicts COVID-19 population adjusted deaths. Below each Moran plot is a loess plot of the main variable (x-axis) in comparison to the categorized variable (y-axis). Moran’s plots for all variables are available in the supplemental materials.

Regression analysis results were performed for the coterminous United States, as well as by region, with full results found in the provided supplemental materials. The *R*^2^ for regional US regression models varied across the differing variant windows (Alpha *R*^2^ = 0.41 (Region 7) to *R*^2^ = 0.83 (Region 9); Delta *R*^2^ = 0.65 (Region 6) to *R*^2^=0.90 (Region 3); Omega *R*^2^=0.47 (Region 7) to *R*^2^ = 0.90 (Region 9). Overall, regional *R*^2^ were moderately higher than national regression model values (US Alpha *R*^2^ = 0.416; US Delta *R*^2^ = 0.675; US Omicron *R*^2^ = 0.505). Across all variant windows, region 10, regions 1 and 2 and region 9 consistently performed best, with region 4, region 8 and region 7 having the lowest *R*^2^ values (Fig. 4). Which predictors were significant varied considerably by region and time window, but several patterns were seen. For example, uninsured adults and housing composition were significant for all time windows for region 6; diabetes was significant across all time windows for region 4; voting was significant for singular time windows for a number of regions, with regions 9 and 10 having two time windows that were significant; and vaccination rate, as a proxy for protective health behaviors, became non-significant across most regions as the pandemic progressed, except for region 5.

**Figure 4.**
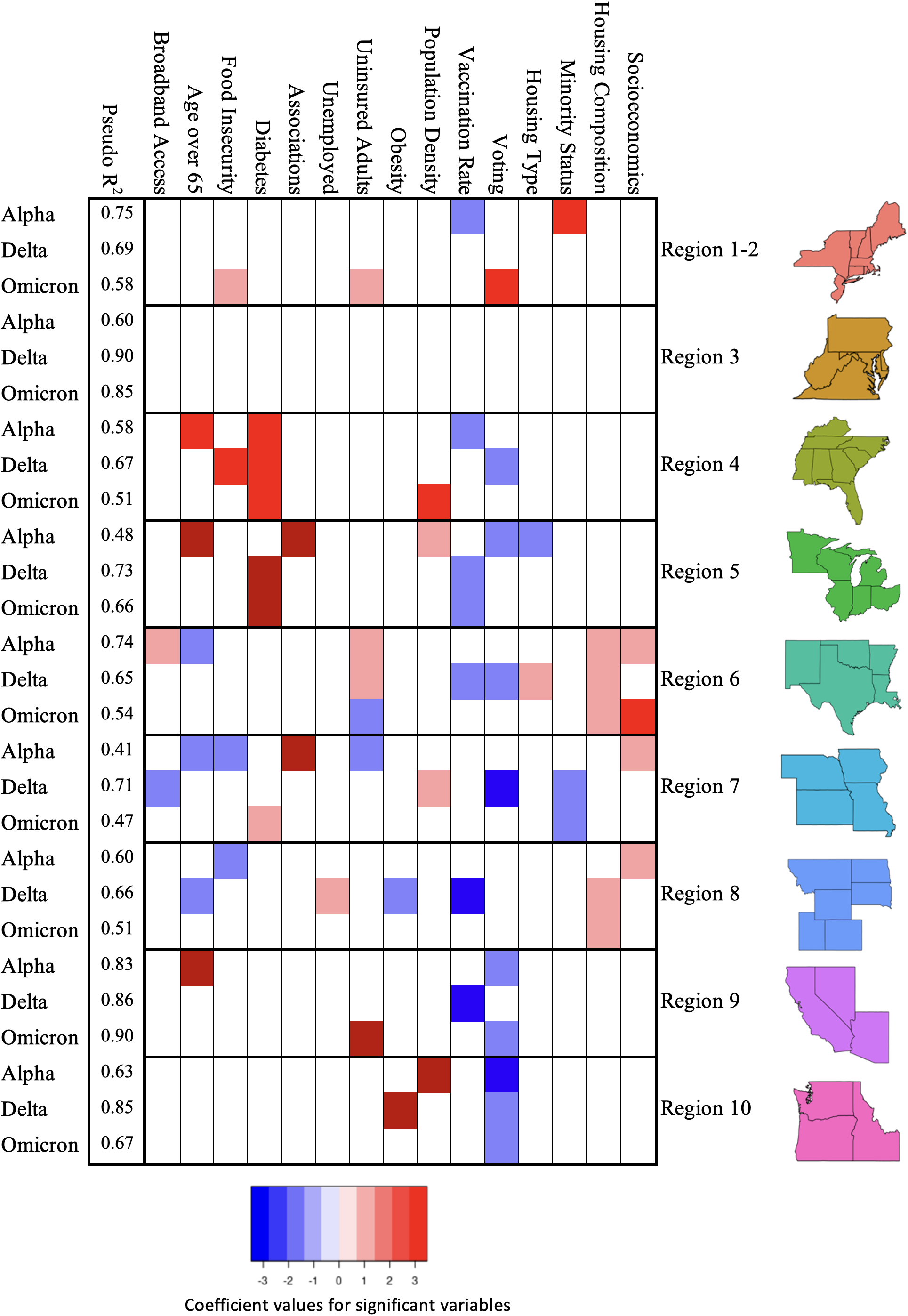
Significant variables for each regional model, as well as *R*^2^ values, per variant time window.

Geographically weighted random forest outputs for the three models in question (Alpha, Delta, Omicron), optimized for bandwidth selection using local/global model root mean square errors (RMSE), performed well in tracking general spatial trends, while overestimating extreme values (Fig. 5). Global out of bag (OOB) *R*^2^ for the three models were 0.40 (Alpha), 0.45 (Delta), and 0.34 (Omicron), while localized *R*^2^ by county indicated a clear variation in predictive power, with values as high as 0.90. The *R*^2^ values in this instance are generated by dividing the vector of mean square errors by the variance of y, then subtracting from 1. Given that the GWRF model approach permutes across a large number (500+) of decision trees based on an adaptive kernel function, there is the possibility of models performing worse than a random outcome (resulting in some *R*^2^ values below zero) [47]. Predictive strength for early in the pandemic (Alpha) was strongest in coastal Eastern regions, the Southwest, as well as Pacific Northwest coastal regions. We see a shift of model performance during the Delta wave, with the west coast regions, the Midwest, and Colorado being highest in terms of predictive power. For Omicron, predictive strength had a more varied spatial distribution, with clusters in Southern California and Arizona. Optimized bandwidth selection (number of counties included in individual county RF model runs) ranged from as high as 24 (Alpha), to 12 (Omicron).

**Figure 5.**
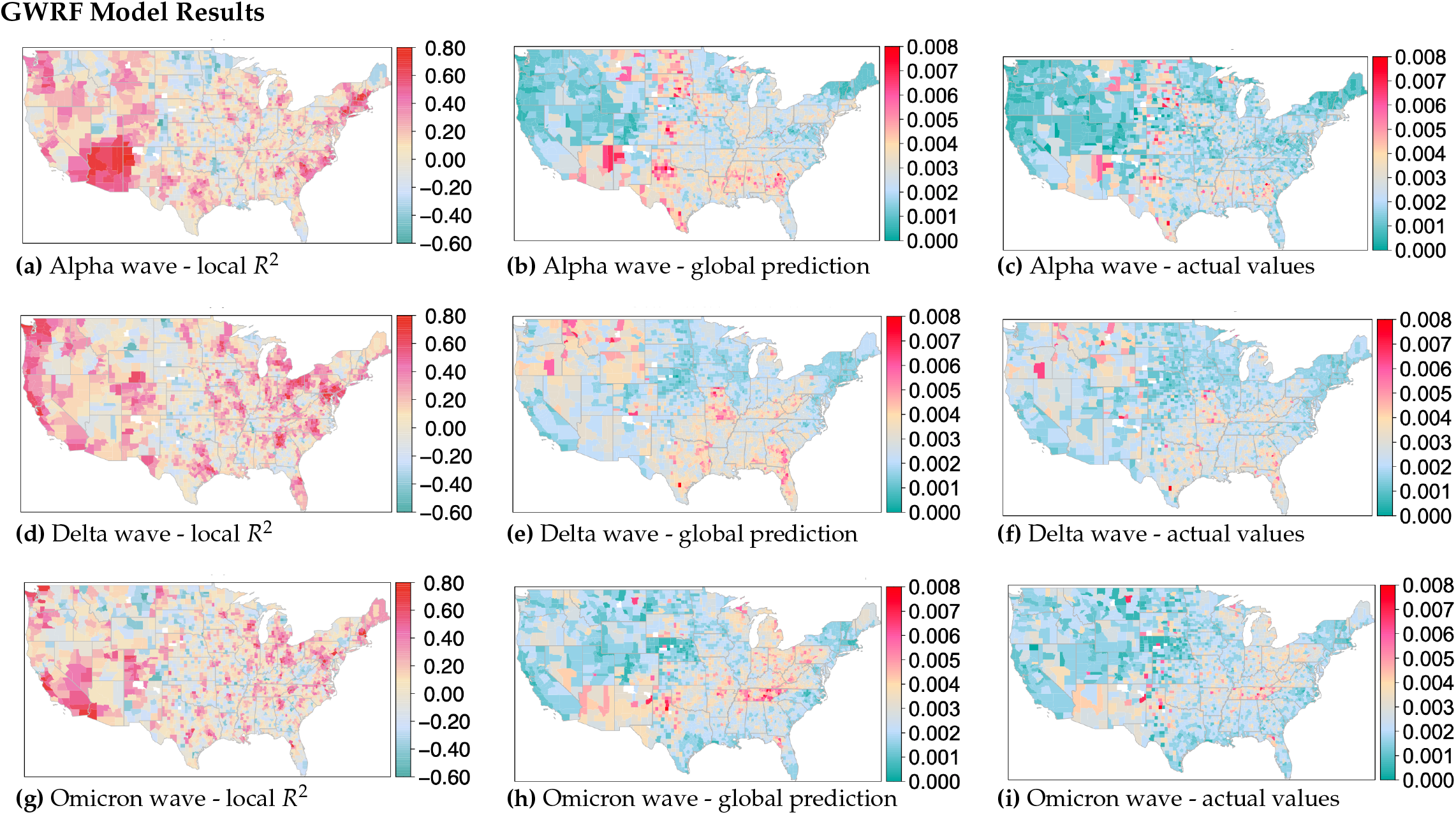
Geographically weighted random forest model results for Alpha, Delta, and Omicron wave time windows. Each panel indicates localized *R*^2^ values, global prediction results, and observed cumulative deaths by county (adjusted for population). a) thru c): Panel plot of Alpha wave results. (c thru f): Panel plot of Delta wave results. g) thru i): Panel plot of Omicron wave results. Full model results for all waves can be seen as part of the supplemental materials.

For GWRF, mapped feature importance values (Fig. 6) provide spatial patterns for predictor variables across all time variant models. For this analysis, feature importance is defined as the average increase in squared residuals of the test dataset, as variables are randomly permuted [42]. Higher values indicate a greater importance of the variable on the model performance. Given this random permutation, there can be instances where the mean square error of a random variable may be higher than a selected variable, generating a negative value. Model results indicate that diabetes feature importances were highly impactful for predicting COVID-19 death in the southwestern portion of the United States across all three time windows; similarly, vaccination rates were impactful in the northern Utah region, across both Alpha and Delta time windows. Household composition influence were high in the Utah/Colorado region for Alpha and Delta time windows as well, while voting was variable across all three variants, with small hotspots in many rural communities. Obesity also provided interesting spatial patterns, with uniquely high feature importance values in the northeastern portions of the United States (Maine). All feature importance plots can be found in the attached supplemental materials (Supplementary Figures S26, S30, and S34).

**Figure 6.**
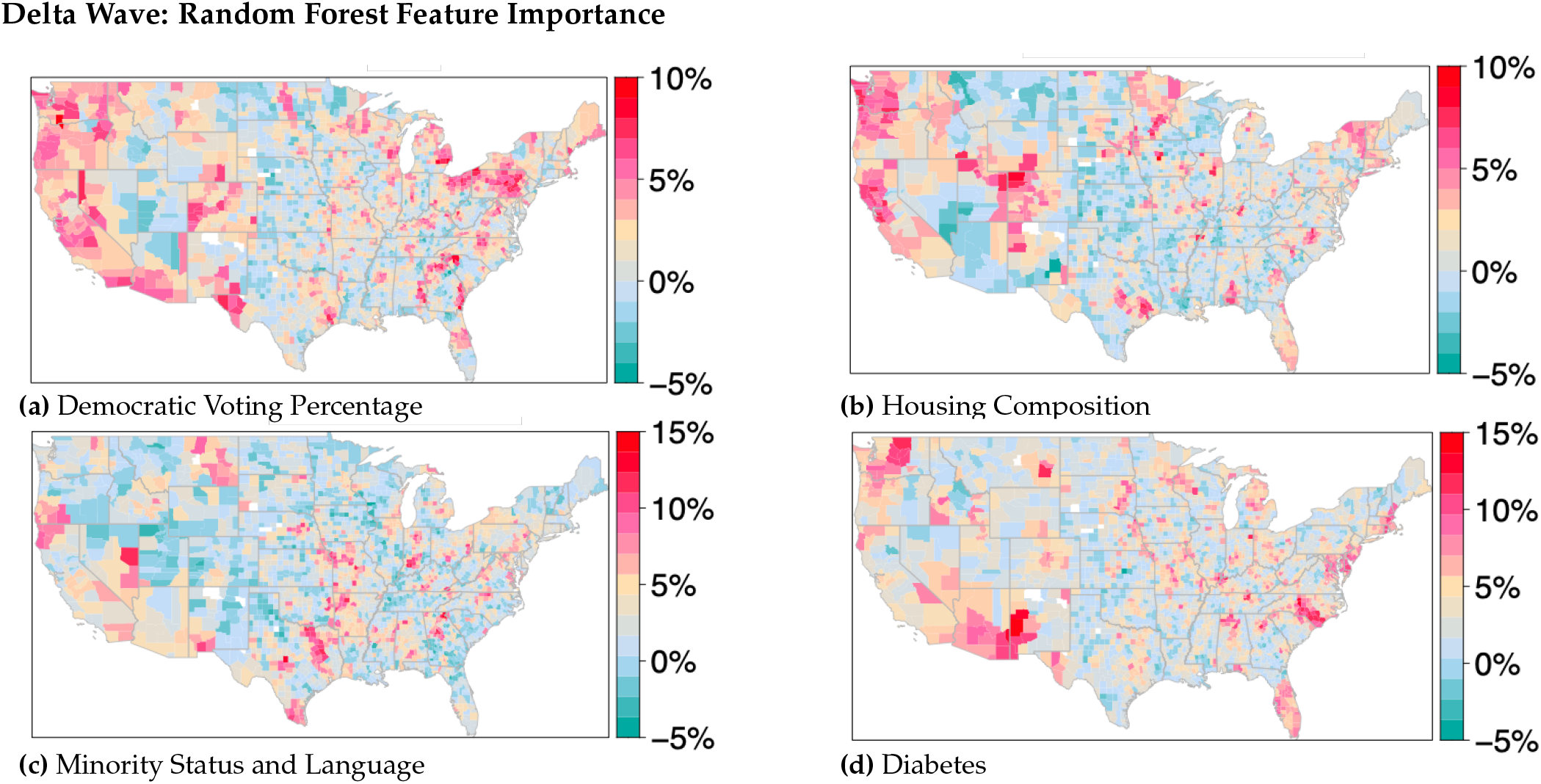
Delta wave random forest feature importance for housing composition, diabetes, minority status and language, and democratic voting percentages (2020 US presidential election). Other feature importance maps can be found in the associated supplemental materials.

Comparisons of regression results with random forest outputs suggest that regional boundaries in the linear model may be limiting predictive capabilities, given its artificial geographic structure. While a number of the regional outputs indicate moderately performing models (region 10 (Pacific Northwest), region 1 and 2 (Northeast), region 9 (West), a majority of the regional regression models had moderate to poor predictive power. Temporal patterns of model performance show higher *R*^2^ values in the Alpha and Delta waves. This suggests that early in the pandemic, typical sociological, economic, political, and health parameters had a greater effect on predicting deaths—yet over time, external effects became more influential. Such effects might include: varying policy response, changing economic factors, social media misinformation, and increasing population immunity either through exposure to the virus, vaccination, or a combination. Conversely, our customized GWRF nationwide model performed considerably better, with feature importance at a county level indicating clustering patterns which do not conform to HHS regional boundaries. Global model results had *R*^2^ values between 0.88 and 0.90. Out of bag *R*^2^ values for GWRF outputs ranged from 0.34 to 0.45, suggesting that, while general trends can be seen, the the model is not able to accurately predict the cumulative deaths for counties in the tails of the distribution. Feature importance rankings displayed interesting spatial patterns (See Supplementary Figs. S26, S30 and S34). Urbanized centers showed higher model influence for minority status and language across all three time windows; socioeconomic measures had the most effect on model performance in rural areas, particularly in the western portions of the United States. Voting showed considerable heterogeneity in terms of model influence, with small pockets of increase in rural regions as well.

## 4. Discussion

The United States recorded over 1.1 million excess deaths during the period of the COVID-19 pandemic from January 2020 through March 2022 [48], with some communities experiencing much greater losses than others. While our regional regression models were able to explain a large portion of the variation in many cases, our geographically weighted approach applied with non-parametric random forest methods better captures variation corresponding with geographic heterogeneity than the regional regression models. For example, in Region 3 not a single variable was significant despite the high *R*^2^ values (Figure 4). Applying spatial modeling methods exposes spatial relationships among predictor variables which are not evident through more traditional modeling techniques which aggregate effects across the modeled region.

Political ideology was identified early in the pandemic as an atypical predictor of deaths due to COVID-19 [49]. Our work supports the finding that political ideology, as measured by voting data, serves as an important feature for predicting variation in COVID-19 deaths in some areas and at certain time intervals, such as the western United States during the Delta wave (Figure 6). Population adjusted deaths for all three time frames show positive spatial autocorrelation—with conservative voting from the 2020 general election aligned with higher deaths, and democratic voting aligned with lower deaths (Fig. 3a). This reinforces previous research which indicates that stronger exposure to conservatism corresponds to higher age-standardized COVID-19 mortality rates [50]. Results from our GWRF modeling also indicate an increasing interaction between political ideology and the changing COVID-19 climate over time as the feature importance of voting increased for the Delta and Omicron waves (Figure 6). This effect, occurring after many local public health restrictions were lifted and the vaccines were widely available, was especially notable in areas of the western and northeastern United States.

We expected that measures of obesity and diabetes would be important for predicting deaths, as these health factors are known to be strongly associated with increased risk of morbidity and mortality from COVID-19 [51]. Indeed, spatial trends for both obesity and diabetes in all waves indicate that increasing spatial autocorrelation corresponds to a higher COVID-19 death rate, as seen for the Delta wave in Figure 3d. However, the only region for which one or both of these variables was significant in the Poisson regression models in all waves was the southeast (Region 4). This demonstrates that important vulnerabilities that may not be aggregated across the region can still be detected by our GWRF feature importance plots.

All of the composite SVI measures, which were constructed for use by FEMA for responding to disasters [52], showed a positive association with deaths in the Moran’s I plots in the first two waves. This relationship was less marked for the Omicron period, with only the measures corresponding to socioeconomic status and minority populations continuing the pattern. This supports previous findings [53,54] that these measures are also useful for predicting clusters of counties with high vulnerability to pandemic threat. Additionally, spatial autocorrelation of both dependent and independent variables show clustering patterns which suggest that particular factors may have differing spatial influences on deaths on whole, see Part 4 of the Supplement. For example, for vaccination rates we see decreasing spatial autocorrelation with increasing deaths, especially in the Delta Wave (Figure S20), while for diabetes we see opposite relationship (Figure 3d). Overall, spatial lag variations between independent variables is reflective of local socioeconomic and cultural views which can dampen (or exacerbate) the effects of COVID-19 associated factors (e.g. deaths) [55].

Select regional analyses provide insights into the value of a spatially explicit modeling approach. For example, variable importance values for obesity across the Delta and Omicron waves show a unique hotspot for the state of Maine. Given Maine’s demographics as older, rural, yet politically progressive, model results suggest that obesity has a uniquely strong influence on predicting COVID-19 associated deaths, which have associations with statewide policy or regulatory components [56]. Comparatively, older populations (over 65) had a moderately stronger influence on model performance in southeastern rural areas (northeastern Georgia, northern Mississippi) across all three variant time windows. This pattern was similar to other regions of the United States, where older populations in rural regions showed higher levels of model influence (central Texas, northeastern Pennsylvania). Such patterns reinforce understandings of rurality on health outcomes, which may be related to poverty and preventative care access [4].

One limitation of the GWRF models is their ability to predict extremes of both high and low levels of deaths. This may be due to confounding variables which are not included in the model which could include, but are not limited to, health parameters (hypertension, cardiovascular disease) and hospitalization effectiveness. Limiting model time windows to the three main variant waves could also be a concern. Future analyses could address these issues by incorporating autoreggressive integrated moving average (ARIMA) techniques, in combination with spatially-focused random forest, or potentially other spatiotemporal algorithms (or an ensemble of multiple algorithmic techniques) [57].

Early in the pandemic, deaths were generally higher in more liberal, population dense regions of the United States. Over time, we see a shift in deaths to more conservative, rural communities, which is likely a combination of vaccination rates and the overall spread of COVID-19 to lesser populated regions. We also see specific regions within the United States (southern Mississippi Delta region, southwestern portions of Arizona, Colorado, and New Mexico) that show variable clustering as well as high levels of model predictive power (Fig. 5). These clusters span multiple HHS boundaries, which helps to explain the weaker regression modeling performance for certain sections of the country. Given spatial and temporal variation of deaths and the inability for linear modeling techniques to effectively perform, policy decisions for future pandemic responses should consider spatially and temporally sensitive modeling efforts to assess public need [29]. In particular, collaboration across boundaries for health regions, which normally operate independently for funding allocations and policy decision, may be necessary for successful interventions. Our GWRF results indicate a considerable difference in spatial feature importance patterns between all three wave events, which align with more qualitative examinations of the pandemic response in the United States.

## Author Contributions

E.S., J.J.L. and C.R.M. conceived the analyses, E.S. conducted the analyses, and E.S., J.J.L. and B.J.R. analysed the results. All authors reviewed the manuscript.

## Data Availability Statement

A supplementary appendix as well as associated datasets can be found at: https://doi.org/10.5061/dryad.4j0zpc8j1

## Acknowledgments

We would like to thank the pandemic modeling group at the Institute for Modeling Collaboration and Innovation (IMCI) at the University of Idaho for help working on and thinking about COVID-19 related issues. Finally, we thank Dr. Holly Wichman for her invaluable research leadership at IMCI.

